# Effective reproduction number for COVID-19 in Aotearoa New Zealand

**DOI:** 10.1101/2020.08.10.20172320

**Authors:** Rachelle N. Binny, Audrey Lustig, Ann Brower, Shaun C. Hendy, Alex James, Matthew Parry, Michael J. Plank, Nicholas Steyn

## Abstract

The effective reproduction number, *R_eff_*, is the average number of secondary cases infected by a primary case, a key measure of the transmission potential for a disease. Compared to many countries, New Zealand has had relatively few COVID-19 cases, many of which were caused by infections acquired overseas. This makes it difficult to use standard methods to estimate *R_eff_*. In this work, we use a stochastic model to simulate COVID-19 spread in New Zealand and report the values of *R_eff_* from simulations that gave best fit to case data. We estimate that New Zealand had an effective reproduction number *R_eff_* = 1.8 for COVID-19 transmission prior to moving into Alert Level 4 on March 25 2020 and that after moving into Alert level 4 this was reduced to *R_eff_* = 0.35. Our estimate *R_eff_* = 1.8 for reproduction number before Alert Level 4, is relatively low compared to other countries. This could be due, in part, to measures put in place in early-to mid-March, including: the cancellation of mass gatherings, the isolation of international arrivals, and employees being encouraged to work from home.

## Introduction

As of 21^st^ May 2020, more than 5 million people have been infected with COVID-19 globally and over 331,000 have died. In New Zealand, border restrictions and strong interventions to maintain physical distancing (Alert Level 4) were implemented early in the outbreak (Alert Level 4 on 25^th^ March, with 205 cases confirmed and probable) and case numbers have remained comparatively low (1503 confirmed and probable cases; 21 deaths as of 21st May). Daily numbers of new cases have declined steadily since 5 ^th^ April and have remained below 10 since 19^th^ April.

Effective reproduction number, *R_eff_*, is a key measure of transmission potential for a disease. It is the average number of secondary cases infected by a primary case. Values of *R_eff_* greater than one indicate a virus is likely to cause an outbreak, while values less than one mean a virus will likely die out. Global estimates place the mean basic reproduction number for COVID-19, for a fully susceptible population in the absence of any control measures, at approximately 2.6 (s.d.=0.54) (Jarvis *et al*. 2020). However, interventions including school closures, population-wide social distancing and case isolation, reduce *R_eff_* to varying degrees.

Numerous methods have been developed for estimating the reproduction number (Obadia *et al*. 2012). A recent study by Binny *et al*. (2020) used a method by Wallinga & Lipsitch (2006) to analyse case data from 25 international locations and estimate reduction in *R_eff_* after strong interventions were implemented. This method calculates *R_eff_* by relating it to the exponential growth rate of daily new local cases via the generation time distribution (assumed to be known and to remain fixed over time). It assumes a randomly mixed population undergoing exponential growth of new local infections arising by community transmission. The method is therefore not suitable for estimation using case data from the very early stages of an outbreak, when daily numbers of new cases are low and stable exponential growth is not fully established (i.e. growth in case numbers is initially somewhat erratic).

In addition, the method cannot distinguish between imported cases (not arising by community transmission, yet still contributing to future community transmission) from local cases (arising from and contributing to community transmission). Without this distinction, and applying the method to both locally-acquired and imported cases, yields artificially elevated estimates for *R_eff_* because the model assumes that all new cases on a given day arose by community transmission alone. Other methods that do account for imported cases have been developed, however these also rely on suitably high case numbers (Thompson *et al*. 2019). New Zealand’s case numbers are too low for this method to produce reliable estimates.

For these reasons, we did not apply these approaches to New Zealand case data. Instead, we use a continuous-time branching model to simulate COVID-19 spread in New Zealand (Plank *et al*. 2020), and report the values of *R_eff_* from simulations that gave best fit to case data. We present an estimate for *R_eff_* before and after implementation of Alert Level 4, that can be used to inform decision-making and parameter choice for models of COVID-19 spread in New Zealand.

## Methods

We obtained daily case data for New Zealand from Ministry of Health, and used internationally imported cases and locally acquired cases (confirmed and probable) for fitting. The stochastic model of Plank *et al*. (2020) was used to simulate COVID-19 spread with the parameter values given in Table 1 (all other parameter values were as used in Plank *et al*., 2020). The model is a continuous-time branching process that uses the internationally imported cases as seed data and simulates the number of new clinical and sub-clinical cases transmitted within New Zealand each day. The model accounts for two broad classes of control interventions: (i) isolation of confirmed cases, reducing onward transmission; (ii) population-wide reduction in contact rates due to restrictions associated with the Alert Level system. The model allows for the delay from infection to symptom onset (Lauer et al., 2020) and from symptom onset to reporting (based on the New Zealand Data) - see Table 1. A full model description is provided in Plank *et al*. (2020), with extensions described in James *et al*. (2020) to account for individual heterogeneity in transmission rates (e.g. super-spreaders) and a proportion of clinical cases that are undetected by testing. We compared the simulated number of reported cases per day (mean of 1000 realisations) to actual reported cases and estimated best-fit values of *R_eff_* by minimising the root-mean-square error of square root-transformed data, over a time window from 10 March to 27 April. Bootstrap confidence intervals (95%) were obtained by re-estimating *R_eff_* for each of 10,000 simulations from the best-fit model.

**Table 1:** Parameters used for model fitting and sensitivity analyses, and their sources. Reproduction number for clinical infections (no case isolation or control) was set to R_clin_ = 3.

| Parameter | Value | Source | Sensitivity analyses |  |  |
| --- | --- | --- | --- | --- | --- |
| | | | Values tested | $R_{eff}$ pre-AL4 | $R_{eff}$ in AL4 |
| Distribution of generation times | $Weibull(\text{scale} = 5.67, \text{shape} = 2.83)$ | Feretti et al (2020) | $Weib(\text{scale} = 9.0, \text{shape} = 2.83)$,<br>$Weib(\text{scale} = 5.67, \text{shape} = 1.5)$ | 1.6, 1.75 | 0.35, 0.35 |
| Distribution of exposure to onset (days) | $T_1 \sim \Gamma(\text{shape} = 5.8, \text{scale} = 0.95)$ | Lauer et al (2020) | - | - | - |
| Distribution of onset to isolation (days) (from data) | $T_2 \sim Exp(\text{mean} = 2.18)$ | Davies et al (2020a) | $T_2 \sim Exp(\text{mean} = 6)$ | 1.95 | 0.05 |
| Distribution from isolation to reporting | $\Gamma(\text{shape} = 1, \text{scale} = 3.48)$ | Fitted to data | $\Gamma(\text{shape} = 1, \text{scale} = 4.70)$ | 1.8 | 0.3 |
| Relative infectiousness of subclinical cases | $R_{sub}/R_{clin} = 50\%$ | Davies et al (2020a) | 20%, 80% | 1.7, 1.75 | 0.4, 0.35 |
| Proportion of subclinical infections | $p_{sub} = 33\%$ | Davies et al (2020b),<br>Byambasuren et al (2020), Lavezzo et al (2020) | 20%, 50% | 1.8, 1.75 | 0.35, 0.35 |
| Relative infectiousness after isolation | $c_{iso} = 65\%$ | Davies et al (2020a) | 30%, 90% | 1.8, 1.65 | 0.4, 0.35 |
| Proportion of clinical cases detected and reported | $p_R = 75\%$ | Price *et al.* (2020) and estimated from limited NZ data | 50%, 90% | 2, 1.6 | 0.4, 0.35 |
| Individual heterogeneity in transmission rate | $Y_K = 0.5$ (i.e. moderate super-spreading) | Endo et al (2020), Lloyd-Smith et al (2005) | 0.1 (high super-spreading), 1 (low super-spreading) | 1.7, 1.8 | 0.35, 0.35 |

## Results

We estimate that New Zealand had an effective reproduction number *R_eff_* = 1.8 [1.44,1.94] prior to moving into Alert Level 4; after moving into Alert level 4 this was reduced to *R_eff_* = 0.35 [0.28,0.44]. These best-fit estimates provided a good match between simulations and data, as can be visualised in Figure 1. Best-fit *R_eff_* estimates were relatively insensitive to changes in other model parameters, namely: the proportion of cases detected, the generation time, onset-to-isolation delay, isolation-to-reporting delay, proportion of sub-clinicals and their infectiousness, relative infectiousness after isolation, and individual heterogeneity in transmission (Table 1).

**Figure 1:**
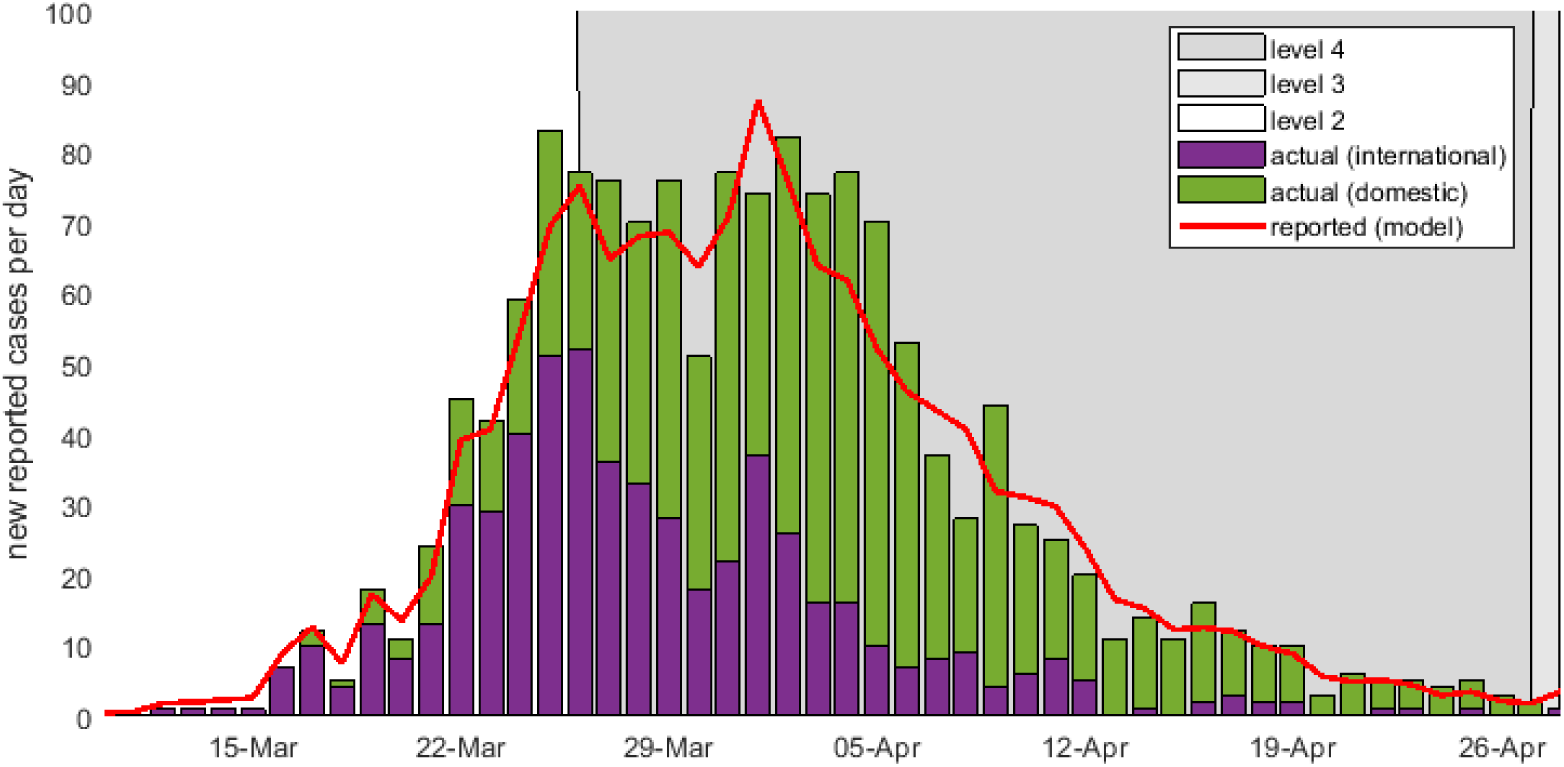
Simulated and actual daily numbers of new local and imported cases (confirmed and probable) in New Zealand prior to and during Alert Level 4. The values of R_eff_ that gave the best model fit to the data, evaluated using a least squares approach, were R_eff_ = 1.8 prior to lockdown and R_eff_ = 0.35 in Alert Level 4.

## Discussion

Our results suggest that New Zealand’s Alert Level 4 was effective at reducing reproduction number to a value less than one, meaning the virus was no longer likely to cause an outbreak for as long as the Alert Level remained in place. The Alert Level 4 estimate *R_eff_ =* 0.35 is an especially small reproduction number and on a par with reproduction numbers reported for other countries whose Alert Level 4-equivalent interventions have been particularly effective (Binny *et al*., 2020). For example, lockdowns in six Australian states have successfully reduced effective reproduction number to approximately 0.3-0.5, and interventions are now being relaxed (Price *et al*., 2020), while other countries (e.g. UK) remain at *R_eff_* ≈ 1 (Binny *et al*., 2020). Our estimate for reproduction number before Alert Level 4, *R_eff_* = 1.8, is relatively low compared to those reported globally. This could be due, in part, to measures put in place in early-to mid-March, for example the cancellation of mass gatherings, fast case isolation of international imported cases, and employees being encouraged to work from home.

We estimated an 81% reduction in *R_eff_* (from 1.8 prior to Alert Level 4, to 0.35 in Alert Level 4) as a result of the Alert Level 4 lockdown. This is in good agreement with estimates from other recent studies, employing a range of different modelling approaches to assess effects of physical distancing on *R_eff_ for* COVID-19. For example, Jarvis *et al*. 2020 reported a 74% reduction in *R_eff_* (from a mean of 2.6 to 0.62), as a result of physical distancing during lockdown in the UK. Their approach used survey data to derive age-structured population contact matrices, to compare the mean numbers of contacts by people in different age-groups, before and during lockdown. Another study by Flaxman *et al*. (2020) fitted a hierarchical Bayesian model to data from 11 European countries to estimate an approximately 50% relative reduction in *R_eff_* due to lockdowns, compared to four other intervention policies. However, most countries the Flaxman *et al*. analysis were still in the early stage of outbreak; so interventions were likely not in place long enough to observe the full extent of their effect. A third study used a Bayesian SEIR-type model to infer a 78% reduction in contacts in British Columbia due to the introduction of physical distancing measures (Anderson *et al*. 2020).

These estimates of reproduction number will be important for informing future decision-making on intervention policies and for improving models of COVID-19 spread in New Zealand. Since de-escalating to Alert Levels 3 and 2, case numbers have been too low to obtain reliable estimates for their *R_eff_* using this approach. It is likely that effective reduction of *R_eff_* under Alert Levels 2-3 *after* Level 4 ‘lockdown’ will differ from the effectiveness of Alert Levels 2 and 3 *before* Level 4 ‘lockdown’ on 25^th^ March. This is because before Level 4, there was high risk of undetected community transmission and growing numbers of local cases; whereas after Level 4 case numbers are low, case isolation is fast, and large clusters have been identified and closely monitored.

## R Calculator and Outbreak Simulator Tools

To leverage the model and results described above, we built an app (http://covid19takecontrol.nectar.auckland.ac.nz/covid19_takeControl/) that allows the general public to interact with the models by ‘driving’ simulated Alert Level decisions and the public’s response to them. COVID 19-Take Control offers a simulator for how case numbers might rise and fall with different levels of public response to Alert Levels of different lengths, an R calculator to compare the relative efficacy of different intervention tools on *R_eff_*, and up-to-date background epidemiological and sociological information.

The app serves two purposes: 1) to communicate the models to the general public in a user-friendly and accessible way; 2) to support New Zealanders in their efforts to make huge changes to their daily lives. As its name implies, COVID 19 – Take Control is premised on prominent cognitive science findings that people are more likely to change their behaviour, even if inconvenient or costly (as lockdowns certainly are), if they feel those individual changes will be effective (Bavel *et al* 2020). Hence the app conveys messages of self-efficacy such as ‘we are the architects of our future.’ Lest those empowerment message seem trite, they are grounded in interactive tools that allow users to see the effect of their actions (in hand-washing, physical distancing, or contact-tracing) on *R_eff_* and on the shape of the epidemic curve. In other words, the app aims to support the general public to recognise the effects of their behaviours, that the ‘power to control COVID 19 is in our hands, quite literally.’

## Data Availability

This article does not present new data. Link to source of data used in analyses provided.

## Acknowledgements

We thank Tom Cunningham for useful discussions on effective reproduction number.

